# Multi-tissue proteomic signatures of Alzheimer’s disease: a systematic investigation in brain, CSF, blood and an exploratory study in tears

**DOI:** 10.1101/2025.01.14.25320538

**Authors:** Jie Shen, Yuhui Huang, Minyu Wu, Minqing Yan, Jing Sun, Hui Chen, Xin Xu, Peige Song, Pang Yao, Xu Qian, Jintai Yu, Xue Li, Tiannan Guo, Changzheng Yuan

## Abstract

Proteomic studies have the potential to identify etiological biomarker and interventional targets for Alzheimer’s disease (AD). However, limited studies have systematically investigated and compared the proteomic profiling related to AD across multiple tissues. First, we systematically reviewed 112 proteomic studies of AD (comprising 107 case-control studies of 16,997 individuals and 5 prospective cohort studies of 60,782 individuals) and synthesized a map of 902 brain bulk, 315 cerebrospinal fluid (CSF), and 9 blood markers that were consistently altered in AD individuals across at least 5 studies. In particular, a total of 55 common proteins altered in the same direction in brain bulk and CSF, whereas 33 proteins altered in the opposite direction. Next, we applied proteome-wide Mendelian randomization and identified 28 brain, 32 CSF, and 59 plasma genetically predicted proteomic markers associated with AD (all FDR < 0.05). The comparison across multiple tissues uncovered a panel of 20 AD-related proteins altered in at least two human tissues. Overall, a pool of 1,219 multi-tissue high confidence proteins were identified in the two-phase investigation. In the exploratory analysis utilizing a matched case-control study among 79 community-dwelling older adults, we detected a total of 845 high confidence protein markers in tears, of which 312 markers altered in a severity-dependent manner across normal cognition controls, mild cognitive impairment (MCI) and dementia stages. Among these, levels of STXBP1, UBE2V1, PALM, PYGB, ST13, and GPD1 were significantly different in dementia or MCI individuals compared to controls (all P < 0.05). Overall, our study provides a comprehensive roadmap of multi-tissue proteomic biomarkers associated with AD to help enhance the exploration of underlying mechanisms, facilitate the development of minimally invasive screening methods, and identify potential targets for novel interventions.

## Introduction

Alzheimer’s disease (AD) is a complex and progressive neurodegenerative disorder characterized by the accumulation of amyloid plaques and neurofibrillary tangles in the brain[1]. Research into potential biomarkers for diagnosis and mechanism of AD has primarily concentrated on key evidence related to amyloid, tau proteins, and APOE gene [2]. The advance of high-throughput proteomics has largely enhanced the ability to identify alterations in protein expression and composition linked to various stages of AD. These proteomic investigations can provide valuable insights into the underlying biological pathways and help identity novel biomarkers for early diagnosis and therapeutic targets across the disease’s progression.

Recent developments have facilitated extensive proteomic profiling of AD across brain tissues[3], including both bulk regions and specific neuropathological lesions, as well as in cerebrospinal fluid (CSF)[4] and blood samples [5]. Although many novel protein biomarkers have been identified in observational studies, definitive evidence remains elusive due to issues including multiplatform challenges, limited sample sizes, and inconsistencies in proteome coverage and reproducibility. Moreover, tissue-specific protein changes have not been thoroughly investigated, hindering deeper mechanistic understanding. In parallel, emerging proteomic studies of urine and saliva are beginning to reveal potential non-invasive biomarkers, although these sample tissues generally reflect short-term changes and may not fully capture long-term brain alterations. Thus, robust evidence for reliably guiding the screening, validation, and diagnostic application of protein candidates in large-scale studies remains lacking.

The connection between the eye and the brain is increasingly recognized in both experimental and clinical therapies[6]. In light of this relationship, several well-defined neurodegenerative diseases, including AD and Parkinson’s disease, which impact the brain and spinal cord, also have manifestations in the eye, with ocular symptoms often preceding neurological symptoms [7]. Notably, postmortem examinations of AD patients have found retinal Aβ plaques [8], and analyses of tear fluid have shown elevated levels of Aβ and tau proteins in individuals with mild cognitive impairment and AD dementia [9–11]. Therefore, the non-invasive nature of tear fluid collection, combined with its unique biochemical profile, could serve as a cost-effective method for biomarker discovery. By building on established biomarkers found in the brain and CSF, further investigation into the proteomic profile of tear fluid may offer crucial insights into the progression of AD, advance early diagnostic capabilities, and enable timely interventions.

This study utilized a multi-faceted approach to investigate proteomic biomarkers related to AD across multiple human tissues. In the first phase, we conducted a systematic review of high-throughput proteomic studies of AD in brain bulk, CSF, and blood, compiling a map of high-confidence candidates for AD staging while addressing issues of inter-study consistency. In the second phase, we performed a proteome-wide Mendelian randomization (MR) analysis leveraging published multi-tissue genome-wide association studies (GWAS) data, aiming to identify causal protein biomarkers of AD. By integrating these findings, we interpreted tissue-specific and common protein expression profiles across different tissues. In an exploratory analysis, we conducted a tear proteomic case-control study to test the presence of the identified high-confidence candidates in tear fluid. We further identified promising proteins linked to dementia progression stages, deepening our understanding of non-invasive proxy for detecting protein changes in AD. The comprehensive results from proteomic studies across various tissues, along with the detection of high-confidence protein markers in tear fluid, provide an integrated view for future exploration of mechanisms, development of non-invasive screening methods, and identification of novel intervention targets.

## Results

### Systematic review (SR) phase

#### Characteristics of included studies

A total of 3,255 articles were included in the initial search (Searching strategy see **Table S1**). After title/abstract screening and full-text review, 112 proteomics studies fulfilled inclusion criteria for integrated investigation of multi-tissue proteomic signatures of AD (**Figure 1A** and **Table S2**). Specifically, 50, 36, and 26 proteomic studies examined the differentially expressed proteins (DEPs) between AD and controls in bran bulk, CSF, and blood tissues, respectively. Some studies examined multiple human tissues, resulting in more comparisons than the total number of publications, as each instance was counted as a distinct comparison. The sum of multi-tissue comparisons exceeded the number of included publications due to some studies examining multiple human tissue, with each instance counted as a distinct comparison in our analysis. In sum, we integrated 7,549, 4,853, and 2,631 AD DEPs across brain bulk, CSF, and blood tissues, respectively, using a multiple testing corrections threshold of 5% in each study.

**Figure 1.**
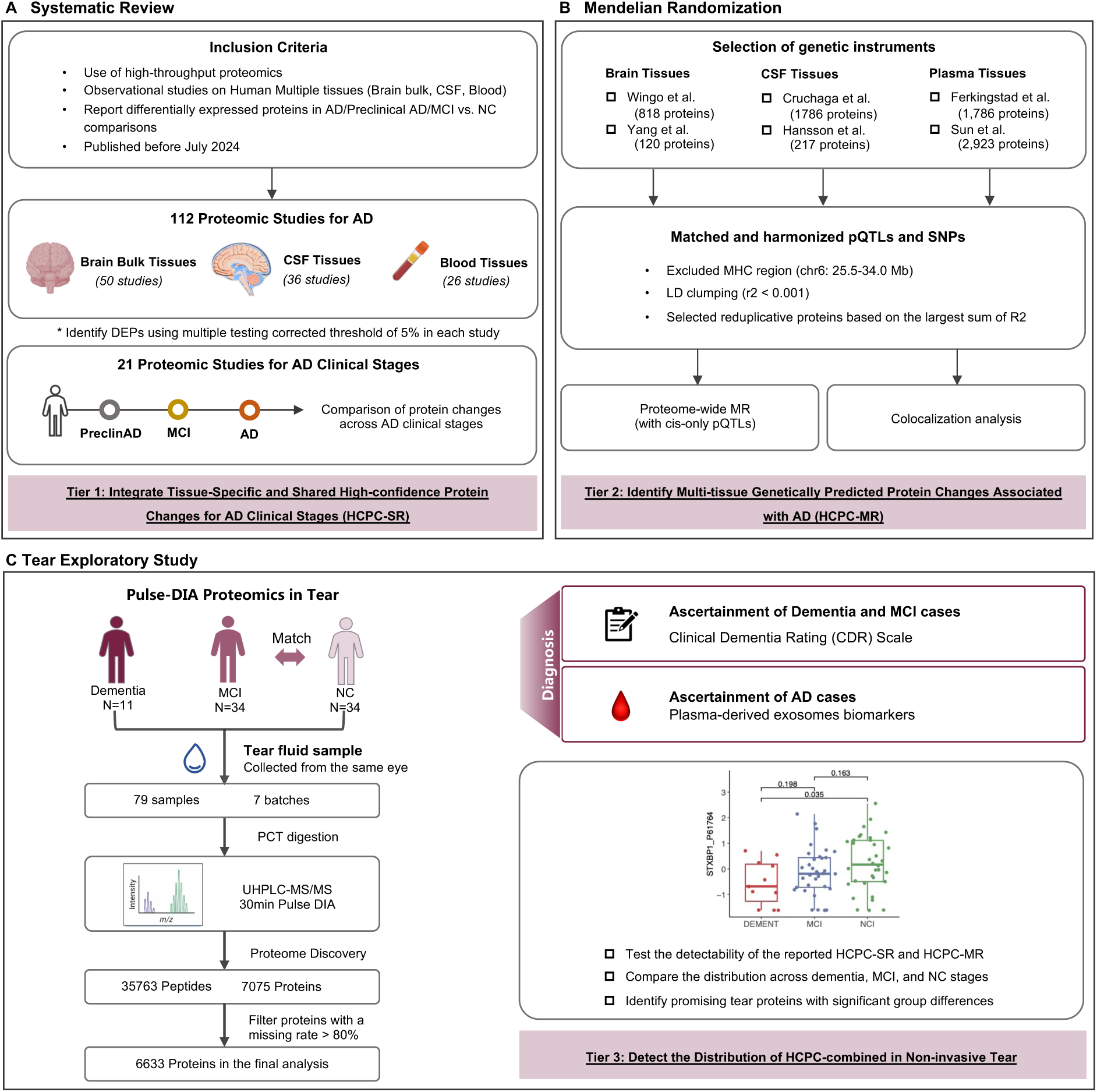
Schematic of integration workflow of multi-tissue proteomic studies for biomarker analyses in Alzheimer’s disease used in current study The study included systematic review, mendelian randomization analysis, and tear proteomic exploratory study to identify and integrate the AD associated proteins for multiple human tissues. **A** We performed a systematic literature review to identify high confidence AD DEPs from published brain bulk, CSF, and blood proteomics, with a focus on ensuring high inter-study consistency (Phase A). **B** Second, Mendelian Randomization analyses was conducted to evaluate causal effects of genetically predicted proteins levels on AD (Phase B). **C** Utilizing the pulse-DIA proteomic analysis, we test the detectability of high confidence AD DEPs in tear fluid sample and propose promising proteins for dementia progression (Phase C).

#### Tissue-specific high-confidence protein changes in AD

In the tissue-specific analysis, we identified 902, 315, 9 protein changes in AD across brain bulk, CSF, and blood tissues as high-confidence protein changes in SR phase (HCPC-SR), respectively, with the criterion being AD DEPs in ≥5 different studies of specific tissues and consistent alteration in the same direction (**Figure 2A** and **Table S3-5**). For brain bulk tissues, 332 out of 920 proteins exhibited consistently increased expression in AD compared to controls, while 570 proteins displayed decreased levels. Additionally, 164 brain bulk proteins displayed inconsistent directional changes in AD. For CSF tissues, 283 out of 315 proteins exhibited consistent increases in Alzheimer’s disease, while 32 proteins showed decreases. Despite this, 48 CSF proteins were characterized by inconsistent expression. For blood tissues, only 9 proteins were consistently altered in AD (8 increased and 1 decreased) and 2 proteins displayed inconsistent changes. These proteins demonstrated a remarkable inter-study consistency in the direction of expression changes across different proteomics studies. Noteworthy is that inconsistency between studies may not attributed to technical issues but could instead significant pathological protein differences across specific disease stages or within distinct tissue regions, such as different areas of brain bulk.

**Figure 2.**
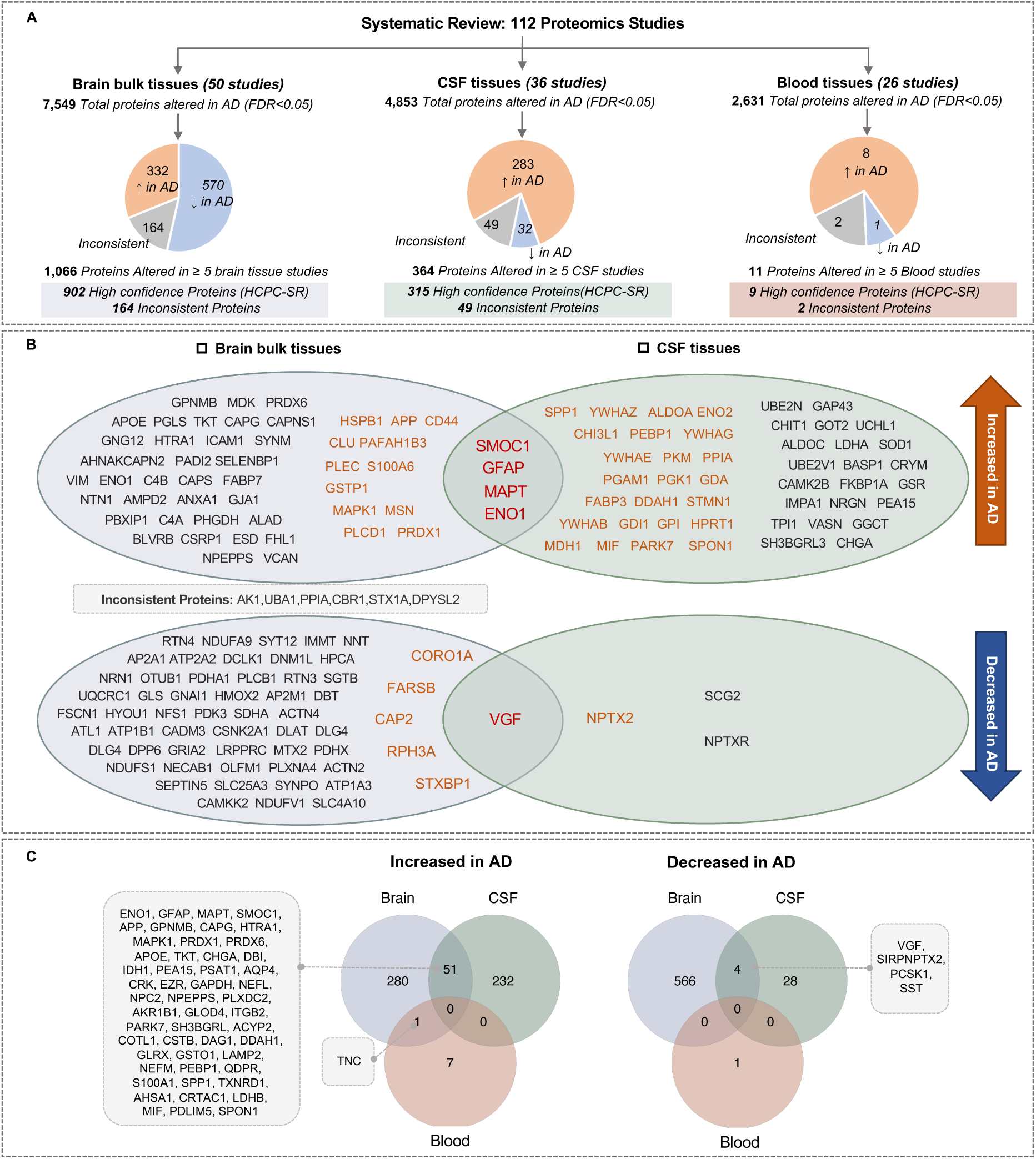
High confidence protein changes with consistent direction in AD across multiple human tissues **A** Breakdown of the number of protein differences in AD among 112 published proteomic studies, after quantification at a 5% false discovery rate (FDR) or Bonferroni comparisons for AD DEP identification in each study. In the systematic review phase, a total of 1,066, 364, and 11 high-confidence proteins that altered in ≥5 brain bulk, CSF, blood tissues proteomic studies, respectively, as shown in pie chart. **B** Most common high confidence proteins across bran bulk and CSF tissues, defined as consistently altered in ≥10 studies corresponding to each tissue type. In brain bulk tissues, 113 most common protein changes in AD, among which 107 were consistently increased/decreased, 6 were inconsistently altered with more than one outlier. In CSF tissues, 54 most common protein changes in AD and 0 proteins inconsistent altered. Blood tissue is not displayed, as all AD proteins were consistently altered in <10 studies. Text colour indicates whether proteins are consistently altered in ≥13 studies (Orange) or in 10-12 studies (Black). **C** Shared AD high confidence candidates within consistent directional change between multiple tissues. 55 proteins altered consistently between brain bulk tissue and CSF, while 1 protein altered consistently between brain bulk and blood. No proteins displayed consistent alteration between CSF and blood.

Specifically, brain bulk proteomic studies summarized 107 most common HCPC-SR which identified as DEPs of AD in at least 10 studies and consistently altered in the same direction (**Figure 2B** and **Table S3**). Of these, 50 proteins were consistently increased in AD and 57 were consistently decreased in AD. The most consistently increased proteins in AD within human brain bulk tissues were predominantly GFAP, HSPB1, MAPT, PAFAH1B3, APP, PLEC, CS44, S100A6, GSTP1, MAPK1, MSN, PLCD1, PRDX1, CLU, and SMOC1, while the most consistently decreased proteins were STXBP1, FARSB, CAP2, RPH3A, VGF, and CORO1A (consistent regulation in ≥13 studies). Additional 6 proteins were inconsistently altered, including AK1, UBA1, PPIA, CBR1, STX1A, and DPYSL2.

In CSF tissue, we compiled 54 most common HCPC-SR which identified as AD DEPs with same directional changes in at least 10 studies (**Figure 2B** and **Table S4**). Of these, 50 proteins were consistently upregulated in AD, with SPP1, YWHAZ, ALDOA, CHI3L1, ENO2, PEBP1, SMOC1, YWHAG, DDAH1, PGAM1, PGK1, PKM, PPIA, and YWHAE being the most prominent (consistent regulation in ≥15 studies). Conversely, 3 proteins - NPTX2, NPTXR, and VGF - were consistently downregulated in AD. Notably, no proteins exhibited inconsistent expression directions among most common CSF candidates.

However, only a small set of HCPC-SR present in 5 or 6 different studies involving blood tissues, including VCAM1, A2M, AMBP, HP, IGFBP2, MMP9, SERPINA1, and TNC, which were consistently upregulated, and PTPN6, which was consistently downregulated (**Table S5**).

#### Shared high-confidence protein changes across different human tissues

We then assessed the consistency or disparity of HCPC-SR across three human tissues. Among the subsets of 902, 315, and 9 HCPC-SR specific to brain bulk, CSF, and blood tissues, respectively, we identified 88 protein changes that occurred in both brain bulk and CSF tissues, 1 common to both brain and blood tissues, while no protein changes that were shared between CSF and blood tissues (**Figure 2C**, **Figure S2A** and **Table S6**). In the comparison between brain bulk and CSF, 55 out of 88 proteins were consistently altered in the same direction. Of these, GFAP, MAPT, APP, MARPK1, SMOC1, PRDX1, APOE, PRDX6, ENO1, and NEFL were predominantly upregulated in AD, while VGF were predominantly downregulated in AD. 33 out of 88 proteins were inconsistently altered between brain and CSF tissues. Additionally, only TNC shared a consistent elevated expression in AD between brain bulk and blood tissues.

#### Complication of protein changes in early stages of AD

We analyze 20 proteomic studies reported protein differences in early stages of AD across three human tissues, consisting of 11 studies of preclinical AD and 13 studies of MCI. For CSF, 59 and 28 proteins were consistently altered in preclinical AD and MCI, respectively, with the criteria of being identified in 2 or more studies with FDR correction of 5% (**Figure S3** and **Table S7**). Based on 315 HCPC-SR for CSF, we observed 27 proteins that consistently altered in the same direction in preclinical AD, and another 27 proteins consistent directional changes in MCI. Notably, SMOC1, DDAH1, GAP43, and GGCT emerged as promising early biomarkers for AD clinical staging, showing the consistent evaluated expression across preclinical AD, MCI, and AD stages.

However, the proteomic data examining protein changes in early stages of AD within brain bulk and blood tissues is less comprehensive, with the formal reporting 2 proteins - HSPB1 and AGRN - consistently increased in preclinical AD in 2 or more studies.

### Proteome-wide Mendelian randomization phase

#### Tissue-specific genetically predicted candidates in AD

The study design of proteome-wide MR phase is shown in **Figure 1B**. Briefly, we employed protein quantitative trait loci (pQTL) data derived from six large-scale proteomic studies and examined their associations with AD (details see **Table S8**). A total of 820 pQTL for 811 brain proteins, 946 pQTL for 944 CSF proteins, and 2,624 pQTL for 2,370 plasma proteins were remained. Using the Wald ratio or IVW method, genetically predicted levels of 28, 32, and 59 proteins were significantly associated with AD risk specific to brain, CSF, and plasma tissues, respectively, with an FDR-adjusted significance threshold of 5% (**Figure 3A**). These proteins were defined as high-confidence AD protein changes derived from MR phase (HCPC-MR). In brain tissues, 13 out of 28 proteins were associated with increased risk of AD, while 15 proteins were linked to a decreased risk (**Figure 3B**). In CSF tissues, 15 out of 32 proteins were linked to an increased AD risk, while 17 proteins linked to a decreased risk. In plasma tissues, 28 out of 59 proteins were found to be associated with an increased risk of AD, and 31 proteins showing a decreased risk. All results of the proteome-wide MR are shown in **Table S9**. The colocalization analyses were performed and the results are summarized in **Table S10**.

**Figure 3.**
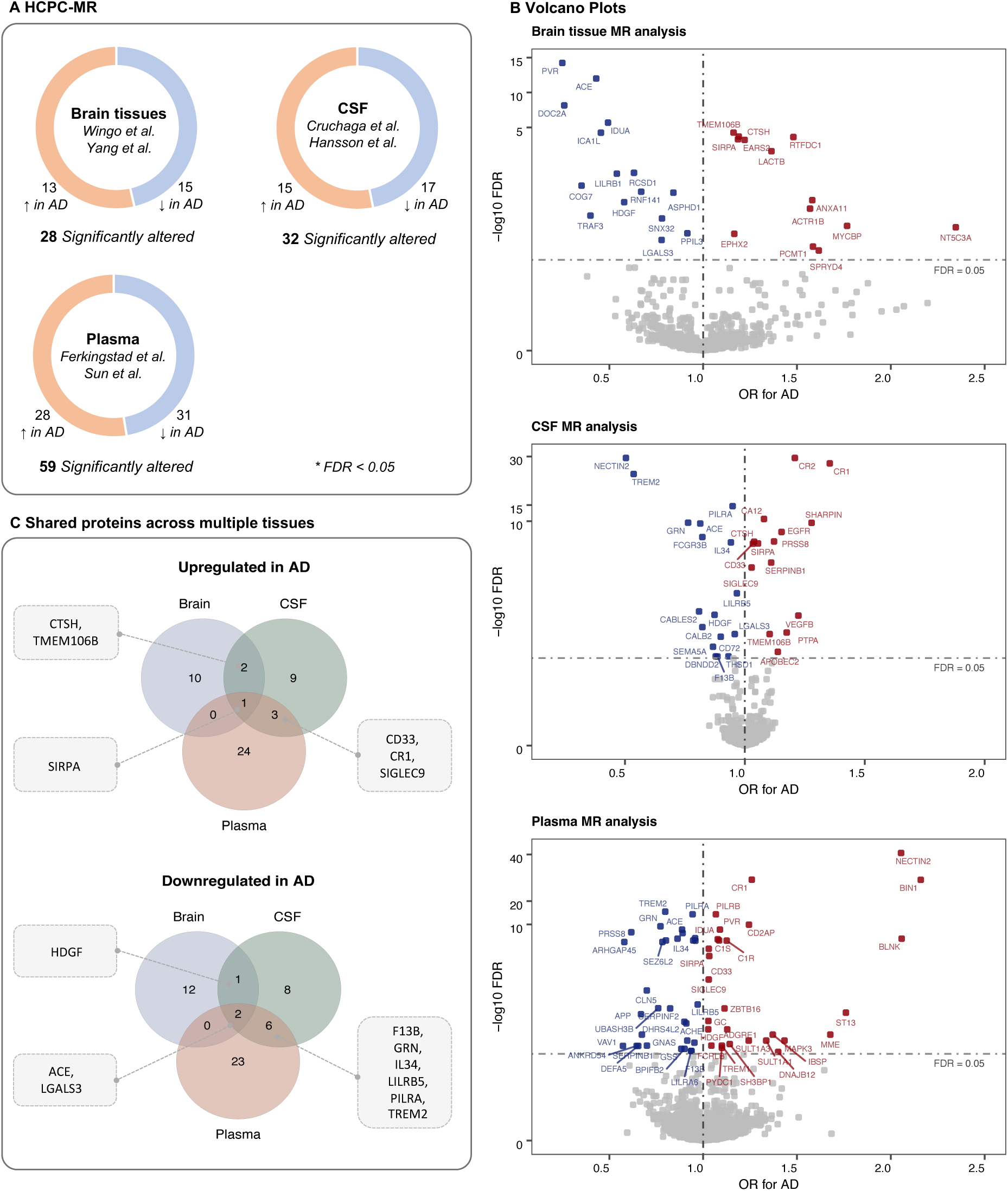
Results from multi-tissue proteome-wide Mendelian Randomization **A** Breakdown of the genetically predicted proteins associated with AD at an FDR-adjusted *P* < 0.05. In the Mendelian Randomization phase, a total of 28, 32, 59 proteins were significantly associated with AD in brain tissue, CSF, and plasma, respectively. **B** Volcano plot showing the results from proteome-wide MR across brain tissues, CSF tissues, and Plasma tissues. X-axis indicates the OR for AD associated with each protein. Y-axis shows the -log10 level FDR value and the dotted line indicates the cutoff of FDR < 0.05. **C** Shared AD associated proteins within consistent directional change between multiple tissues.

#### Shared genetically predicted proteins across different human tissues

The comparison across multiple tissues uncovered a panel of 20 AD-related proteins altered in at least two human tissues (**Figure S2B**). Genetically predicted higher levels of SIRPA were consistently associated with an increased risk of AD across brain (OR _per SD increase_ = 1.18, 95% CI, 1.10-1.27), CSF (1.06, 95% CI, 1.03-1.08), and blood tissues (1.03, 95% CI, 1.02-1.04), while ACE (0.43, 95% CI, 0.34-0.53 for brain; 0.81, 95% CI, 0.77-0.86 for CSF; 0.89, 95% CI, 0.86-0.92 for blood) and LGALS3 (0.77, 95% CI, 0.67-0.9 for brain; 0.96, 95% CI, 0.93-0.98 for CSF; 0.92, 95% CI, 0.87-0.96 for blood) shared an inverse association with AD risk (all FDR < 0.05, **Figure 3C**). Higher HDGF protein level in brain was associated with reduced risk of AD (OR _per SD increase_ = 0.57, 95% CI, 0.43-0.76) and CSF (0.87, 95% CI, 0.81, 0.94), whereas its elevated level in plasma showed an increased association with AD risk (1.03, 95% CI, 1.01, 1.04) (all FDR < 0.05). CTSH and TMEM106B were consistently associated with an increased risk of AD across brain (OR _per SD increase_ = 1.18, 95% CI, 1.10-1.27 for CTSH; 1.16, 95% CI, 1.09-1.23 for TMEM106B; all FDR < 0.05) and CSF tissues (1.04, 95% CI, 1.02-1.06 for CTSH; 1.10, 95% CI, 1.04-1.17 for TMEM106B; all FDR < 0.05). In the comparison of CSF and plasma MR results, 12 proteins were significantly associated with AD risk: 3 (CD33, CR1, SIGLEC9) consistently linked to an increased risk, 6 (i.e., F13B, GNR, IL34, LILRB5, PILRA, TREM2) to a decreased risk, and 3 (i.e., NECTIN2, PRSS8, SERPINB1) exhibited inconsistent associations between the two tissues.

#### Enriched biological pathway and networks

Our analysis identified tissue-specific protein-enriched pathways that may contribute to biological pathology of AD. **Figure S4** and **Figure S5** show the protein-protein interaction enrichment of 902 and 315 HCPC-SR specific to brain bulk and CSF. There was evidence of significantly decreased synaptic proteins, vesicle proteins, and mitochondrial proteins in brain bulk tissues. However, a significant enrichment of increased proteins associated with particular cellular components or biological processes were found in CSF, including synaptic-vesicle function and mitochondrial proteins. In addition, ribosomal proteins were only enriched in brain tissues that can pathologically impair translation.

Combined the identified high-confidence protein changes (HCPC) in SR and MR phases, the top 15 enriched terms of AD candidates in brain bulk tissues and CSF are presented in **Figure S6**. In brain bulk candidates, pathways enriched in downregulated proteins were primarily neuronal synapse, vesicle recycling/transportation, mitochondrial function, homeostasis, and oxidative processes. The upregulated proteins enriched in pathways involving function, lumen, extracellular matrix, lysosomal activity, nervous system development, and metabolic and catabolic processes. All results of GO enrichment analysis shown in **Table S11** and **Table S12**.

#### Exploratory analysis: Detectability and identification in tear proteomics

To quantify, replicate and detect multi-tissue AD candidates, we performed untargeted pulse-DIA based proteomic analysis of tear samples from 79 older adults (mean age: 73.3 years old) (**Table S13**). The experimental workflow is summarized in **Figure 1C**, quantifying 7,075 tear proteins from 35,763 unique peptides. After filtering out proteins with a missing rate higher than 80%, 6633 tear proteins remained for subsequent analysis.

Among HCPC filtering from SR and MR phases, 811/930 brain bulk proteins, 250/344 CSF proteins, and 46/68 blood proteins have been detected and quantified in tear samples, respectively (**Figure 4A**). The combination of HCPC in SR and MR less than the direct sum of HCPC-SR and CHPC-MR due to some proteins being replicated across two phases. The distribution of most common HCPC presented in tear proteomic study was shown in **Figure S5**.

**Figure 4.**
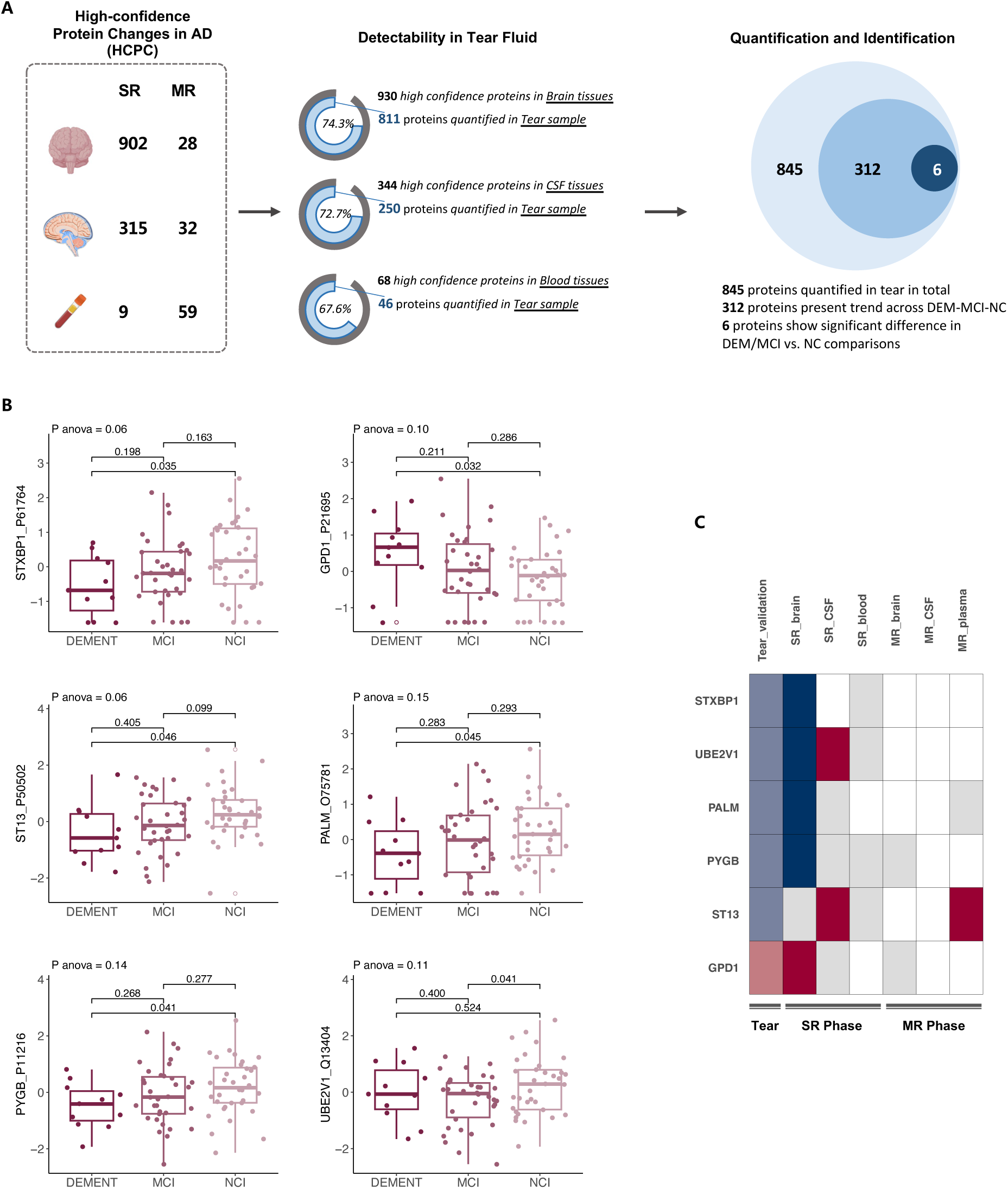
Protein detection and comparison in the tear proteomic analysis **A** Detectability of multi-tissue derived high confidence AD proteins in tear fluid. Based on the integrated multi-tissue AD candidates reported in SR and MR phases, the pulse-DIA proteomic study detected and quantified 845 unique proteins in tear fluid sample. Specifically, 74.3% brain proteins, 72.7% CSF proteins, 67.6% blood proteins have been detected in tear. Among them, 312 proteins increased or decreased in a gradient in MCI and dementia participants. 6 proteins exhibited significant differences in dementia or MCI group (*P* value < 0.05). **B** Expression level change of the 6 promising tear proteins with significant differences between dementia, MCI and CN participants. **C** Heatmap showing the directional change of proteins across brain, CSF, blood (in both SR and MR), and tear (in exploratory study). Colour indicates whether proteins are consistently increased (red), consistently decreased (blue), understudied (grey, defined as <5 publications in SR / FDR > 0.05 in MR). Meanwhile, dark color means high evidence results from SR and MR phases, and light color means replication in the current tear exploratory study.

Of 845 replicated proteins in tear proteomic analysis in total, the median abundance of 312 proteins increased or decreased in a gradient across dementia, MCI, and NC stages. In particular, the abundances of STXBP1, UBE2V1, PALM, PYGB, ST13 were significantly lower in dementia or MCI compared to those in controls, and GPD1 showed increased expression in case groups (P < 0.05) (**Figure 4B**, **Table S14**). Intriguingly, STXBP1, UBE2V1, PALM, PYGB, and GPD1 were also found to be enriched in AD brain bulk, suggesting a potential similarity in protein profiles between tear and brain bulk tissues (**Figure 3C**). Of these proteins, UBE2V1 exhibited opposite directional changes among CSF proteomic studies. The findings suggest that AD progression may also be detectable in some eye-specific pathologies, positioning tear fluid as a valuable non-invasive tool.

## Discussion

Through the systematic investigation of high-throughput proteomic studies involving multiple issues, this study demonstrated the high level of consistency across various proteomic studies related to AD in different tissues. Collectively, we compiled a comprehensive profile of high-confidence protein changes in AD, which includes 930 brain proteins, 344 CSF proteins, and 68 blood proteins, integrating evidence from both systematic review and Mendelian randomization analysis. Enrichments were identified for synaptic, mitochondrial, vesicle recycling, and metabolic pathways within AD brain and CSF tissues. Our findings on tissue-specific and common protein markers across these tissues provide valuable insight into protein alterations associated with AD, and highlights proteins warranting further investigation. Nevertheless, the heterogeneity observed in blood-derived proteins related to AD necessitates careful interpretation and consideration in future research.

Through an exploratory proteomic analysis of tears, we identified potential biomarkers associated with dementia and mild cognitive impairment, including STXBP1, UBE2V1, PALM, PYGB, ST13, and GPD1. Notably, the changes in most tear proteins aligned closely with those observed in brain tissues, indicating their potential as promising non-invasive proxies for early AD screening and diagnostic targets.

This study systematically reviewed and synthesized existing proteomic studies of AD across human brain tissues and biofluids. In appreciation of the similarities and interrelationships between human tissues, the most convincing evidence from the systematic review identified 55 high-confidence proteins with consistent directional changes between brain and CSF, with GFAP, MAPT, APP, MARPK1, SMOC1, PRDX1, APOE, PRDX6, ENO1, and NEFL predominantly upregulated, and VGF downregulated in AD. Previous studies have also revealed that approximately 70% of the CSF proteome overlaps with the brain proteome[12]. The proximity of CSF to the brain offers strong support for deciphering the role of protein biomarkers that linked to AD pathology in both tissues. Potential mechanisms may explain such shared proteins. For example, the APP (amyloid precursor protein) serves as the precursor to Aβ peptides, with APP mutations affecting Aβ cleavage and aggregation[2]. Most mutations in APP alter its processing, resulting in an increased Aβ42/Aβ40 ratio, a hallmark feature observed in AD patients[1]. Similarly, extensive cell culture studies have established APOE (Apolipoprotein E) as a pivotal factor in AD pathology. The APOE functions as a chaperone for Aβ, influencing its clearance and deposition, ultimately contributing to plaque formation [13]. In addition, the MAPT (Microtubule-associated protein tau) represents another pathological hallmark of AD, exhibiting a crucial role of mitochondrial bioenergetic deficiencies in contributing to tauopathy-related autophagy defects[14]. Furthermore, emerging proteome-wide association studies based on human tissues have underscored the significant potential of GFAP (Glial fibrillary acidic protein) in predicting incident AD and dementia[15].

Our findings deepen the understanding of shared alterations in these pre-neuropathological proteins, suggesting their promise for early screening and mechanism research of AD in both brain and CSF. The presence of tissue-specific protein alterations highlights the need for targeted investigations into AD mechanism and therapeutic intervention. Notably, 33 high-confidence AD-related protein markers were found to exhibit opposite expression patterns between brain bulk and CSF tissues. Taking 14-3-3 family proteins as an example, we identified that certain proteins (YWHAZ, YWHAG, YWHAE, YWHAB, YWHAH, YWHAQ) were consistently higher in CSF across multiple studies, many of them showed decreased levels in brain bulk tissues among AD individuals. Numerous studies have revealed that 14-3-3 proteins interact with tau, promoting tau aggregation and/or tau phosphorylation[16,17]. Importantly, recent proteomic studies unveiled that CSF YWHAG and YWHAZ could effectively discriminate AD from normal cognition[4,20].The differential expression between tissues may suggest substantial neuronal damage in the brain tissues and the release of normally retained proteins into the CSF[18].These findings underscore the importance of both consistent and tissue-specific proteins for future diagnostic panel development.

While brain tissue and CSF-based biomarkers have revealed a panel of common proteins, their operational complexity may limit their clinical application. Blood, a minimally invasive and easily accessible sampling method, has better potential for clinical and population-based research. However, evidence on reliable proteomic markers in blood remains less robust. A meta-analysis of 17 independent blood proteomic studies reported inconsistencies in protein regulation associated with AD pathology[5], largely due to relatively crude criteria for identification and no multiple comparison correction restrictions. Building upon established evidence, our systematic review of 26 high-throughput blood proteomic studies identified 9 high-confidence candidates for AD by applying rigorous criteria with multiple comparison corrections and required replication in 5 or more studies. Variations between studies may be attributed to differences in sequencing methods, high false-positive rates from insufficiently stringent criteria, and inconsistent methods for protein identifications. In addition, Mendelian Randomization analyses indicated 59 potential causal proteins, with 31 f genetically predicted proteins (e.g., VAV1, ARHGAP45, PRSS8, SERPINB1, APP) associated with a lower risk of AD, while 28 (such as BIN1, NECTIN2, BLNK, T13, MME) linked to an increased risk. Nevertheless, there is a pressing demand for high-confidence, non-invasive tissue biomarkers that reflect protein changes to facilitate early screening and clinical application.

The connection between the eyes and the brain through the optic nerve, sharing functional neuron and axons, has prompted recent studies into the detection of classical AD biomarkers in tear fluid. Studies have reported elevated levels of Aβ38, Aβ40, Aβ42, total-tau (t-tau), and phosphorylated-tau (p-tau) in patients with MCI and AD dementia[9–11]. The current study, applying untargeted pulse-DIA proteomic in tear sample, demonstrated that approximately 75% high-confidence AD proteins identified from the aforementioned tissues can also be detected in tear fluid. Furthermore, a set of proteins (STXBP1, UBE2V1, PALM, PYGB, ST13, and GDP1) h exhibited consistent regulations across individuals with dementia, MCI, and normal cognition. Of these proteins, 5 out of 6 showing alignment with published brain bulk proteomic studies. Taken together, these shared manifestations between brain bulk and tear fluid indicate the potential of tear samples as a valuable tool for understanding brain mechanisms and processes. Interestingly, the enriched pathways of these promising tear proteins are also linked to synaptic and vesicle functions, similar to those observed in brain tissues. For example, STXBP1 (syntaxin binding protein 1), a synaptic vesicle gene, facilitates membrane fusion and exocytotic release[21]. Animal experiments suggest that UBE2V1 (ubiquitin conjugating enzyme E2 V1) is involved in synaptic plasticity and function, possibly impacting the scaffolding properties of postsynaptic density protein 95 (PSD95)[22]. PALM (paralemmin-1), which increases with age, may reflects cytoskeletal protein degradation and instability [23]. Overall, our study points towards a novel, promising, minimally invasive, and cost-effective approach for early AD detection and diagnosis, suggesting that ocular investigations could provide valuable insights into neurodegenerative disease mechanisms.

Recent studies have proposed that AD progression is initiated by lipid-associated inflammation, followed by altered glucose metabolism, mitochondrial and synaptic function and culminating in oxidative stress responses[24]. The current study identified early protein expression changes in preclinical AD and MCI stages, such as SPP1, YWHAZ, YWHAH, GAP43, NRXN1, PTPRD, PDLIM5, CALB2, and DAG1. Consistent with previous laboratory research[25], our findings support the hypothesis that synaptic and vesicle proteins may serve as early triggers for the pathological alternations contributing to AD, with presynaptic protein changes occurring before Aβ and tau accumulation. Moreover, mitochondria function is significantly linked to oxidative stress, cellular homeostasis, and energy defects in AD pathogenesis[26,27]. Previous proteomics studies of brain tissues in pre-neuropathology AD stages have revealed widespread mitochondria damage typically seen in advanced AD[3]. Our review of CSF proteomics also showed elevated levels of several mitochondria-related proteins in preclinical stages of AD. The mitochondrial cascade hypothesis may help explain such phenomenon, proposing that alterations in mitochondrial function could disrupt Aβ homeostasis potentially impacting disease progression throughout adulthood[28]. However, the overall evidence of proteomic markers in early AD stages was limited and inconclusive, falling short of establishing early protein biomarkers. Future investigations should consider inherent limitations, such as larger sample sizes, advanced proteomic analytical techniques, and integration of multi-omics datasets.

The comprehensive methodologies employed in this multi-tissue omics study constitutes a major strength, allowing for a thorough examination of consistency across studies and identification of genetically predicted proteins related to AD. Adhering to rigorous criteria for multiple comparison correction in the identification of differentially expressed proteins provide more stringent evidence. The comparison of different tissues fosters a deeper understanding of shared and unique proteins, offering valuable insights for the mechanism exploration and development of subsequent diagnosis panels. Notably, this is the first study leveraging high-throughput proteomics of tears to assess the detectability of established AD proteins and proposes a panel of promising candidates in tear fluid, positioning them as a potential non-invasive alternative for AD detection. Nevertheless, there are several limitations. First, only a limited number of proteomic studies linked to early stages of AD, specifically MCI and preclinical AD, posing challenges to draw consistent conclusions about early protein changes profiling. Additionally, preclinical AD might include individuals resilient to AD, thus, resulting in inconsistency of findings within studies. Further large-scale proteomic studies are warranted to address these issues. Second, we did not restrict the specific types of samples within each tissue, such as blood tissue, which could include plasma, serum, or EV samples. This may have partially contributed to inter-study variation. Evidence from various tissues is worth exploring to help us better understand the differences in protein expression between tissues. Lastly, the exploratory analysis of tear proteomics, which included samples from dementia, MCI, and matched controls, may not serve as a direct validation of well-studied AD candidates. Nevertheless, clinical and imaging assessments in dementia patients suggest a subset of cases may involve AD, highlighting the potential of tear fluid proteins for further replication and validation studies.

### Conclusion

In conclusion, our study provides a comprehensive overview of multi-tissue high confidence proteomic biomarkers associated with AD. We synthesized both consistent and tissue-specific proteomic markers and highlighted the potential for detecting these high confidence biomarkers in tears samples. The study findings contribute to a deeper understanding of the proteomic landscape associated with AD, facilitating the exploration of underlying mechanisms, the development of minimally invasive screening methods, and the identification of potential targets for novel interventions.

## Methods

### Phase A. Systematic Literature Review and Exclusion Criteria

We conducted a systematic literature search (up to 16 July 2024) in PubMed, EMBASE, and Web of Science databases for proteomic studies of AD clinical staging. As detailed in **Table S1**, search terms included “Alzheimer Disease”, “dementia”, “cognitive impairment”, “brain tissue”, “hippocampus”, “cortex”, “cerebrospinal fluid”, “blood”, “plasma”, “serum”, “proteomics”, and their variants in English. In addition, the reference lists of selected papers and recent reviews were cross checked to identify any articles that might have been missed. The systematic review has been registered on PROSPERO (CRD42024592432).

Two investigators (J.S. and M.W.) independently assessed the literature eligibility, with disagreements resolved by discussion and consensus with a third author (Y.H.). Using a hierarchical approach, we evaluated titles, abstracts, and full texts to identify eligible proteomic studies that defined protein changes in AD progression stages across brain bulk, CSF, and blood tissues. Articles were considered for inclusion in the systematic review (SR) phase if they met the following criteria: (1) population-based observational study; (2) utilization of high-throughput proteomics, not targeted ELISA/Lumines kits or validation studies on several previously reported proteins; (3) protein identification using traditional statistical methods for one comparison within each proteins, rather than application of machine-learning prediction model or genetically instrument variables; (4) not based on redox, structural, peptide-level proteomics study; (5) demonstration of differential protein expressions between AD clinical stages and control; (6) analysis of brain bulk, CSF, or blood tissues; (7) availability of full list of identified differentially expressed proteins (DEPs) along with p-values for subsequent multiple comparison analysis. In cases where results from a single study were published in duplicate papers, we selected the most recent paper; otherwise, we included the ones with the greatest number of cases or total sample sizes or higher quality. In line with current literature, we defined preclinical AD as the presence of amyloid plaques without cognitive impairment[29]. In addition, proteomics studies examining the proteome of AD pathogenesis (e.g., amyloid plaques, neurofibrillary tangles or cerebral amyloid angiopathy) were excluded. Brain bulk tissues included brain regions, such as frontal cortex, hippocampus, parahippocampal cortex, entorhinal cortex, temporal cortex, cingulate gyrus, cerebellum, and etc. CSF tissues encompassed CSF and related extracellular vesicles samples. Blood tissues included plasma, serum, plates, and derived extracellular vesicles samples. **Figure S1** shows the process of study selection.

Data were manually extracted from published articles and their supplementary materials by two investigators (J.S. and M.W.). In each study, differently expressed proteins between AD clinical stages and controls were defined based on multiple testing corrected threshold (e.g., false discovery rates [FDR], Bonferroni corrections) of <5% using ANOVA and an appropriate post hoc test, Kruskal-Walis H-test, t-test, cox proportional hazards model, or regression model. When multiple comparisons between diseases and control groups were not available in the original study, we estimated the FDR-adjusted p-values and fold change differences between AD clinical stages and controls using published data. To facilitate integrated comparisons between studies, we unified a single gene ID and UniProt ID for each reported protein. For cases where multiple gene IDs were provided for a single protein, the first listed gene ID was selected. Duplicate gene IDs within a study were extracted, and isoform information was removed from UniProt IDs.

### High confidence proteins identification in SR phase

#### Consistency of directional changes for AD

For the SR phase, we firstly generated a tissue-specific protein changes in AD and counted the number of studies that a protein was designated as a DEP in AD proteomic studies (**Figure 1A**). Each protein was assigned separate replicate counts for three human tissues. We then designated proteins as high-confidence AD protein changes in SR phase (HCPC-SR) if they exhibited consistent directional changes (either consistently increased or decreased) in ≥5 tissue-specific proteomic studies, with one directional outlier permitted. Proteins that being presence in ≥5 different studies but displaying two or more outlier directional changes were considered as inconsistent proteins in AD.

#### Comparison of protein changes at early stages of AD

To explore protein changes across the clinical stages of AD, we filtered the proteomic studies that reported DEPs early stages of AD, including preclinical AD and MCI, and subsequently calculated corresponding tissue-specific candidate scores. Due to the stringent threshold for multiple testing corrections applied for DEP identification in each study, only a few proteins were retained for downstream analysis. We designated proteins as potential candidates for preclinical AD or MCI if they were consistently altered in the same direction in two or more studies. All other proteins were categorized as inconsistent proteins within corresponding clinical stages of AD.

### Phase B. Proteome-wide Mendelian randomization (MR) analysis

The study design of proteome-wide MR phase is shown in **Figure 1B**. First, we employed protein quantitative trait loci (pQTL) data derived from six large-scale proteomic studies for the selection of genetic instruments. These proteomic studies provided summary statistics of pQTLs associated with levels of brain proteins[30,31], CSF proteins[32,33], plasma proteins[34,35]. Of these, three studies used SOMAscan platform[31,32,34], two studies used Olink platform[33,35], and one study used TMT[30]. GWAS summary statistics data of 111,326 clinically diagnosed/’proxy’ AD cases and 677,663 controls were used[36].

The following criteria were used to select instruments and proteins: (i) SNPs associated with any protein were selected (P < 5×10^−8^); (ii) the SNPs and proteins within the Major Histocompatibility Complex (MHC) region (chr6:25.5–34.0Mb) were excluded due to their complex linkage disequilibrium (LD) structure; (iii) the LD clumping was then conducted to identify independent pQTLs for each protein (r^2^ < 0.001). For reduplicative proteins among studies, the protein with the largest sum of R^2^ was selected. We further classified instruments as *cis*-pQTLs based on the following criteria: a pQTL was defined as *cis*-pQTLs when the leading SNP in the region was located within 1 Mb of the transcription start site of the protein-coding gene. Finally, a total of 841 (841 cis pQTLs), 1050 (1052 cis pQTLs), and 3281 (3519 cis pQTLs) unique brain tissue, CSF, and plasma proteins were included in the analysis. Instrument variables are presented in **Table S8**.

The “TwoSampleMR” package was employed to perform MR analysis, examining the associations between genetic instruments and AD. For any proteins with only one instrument, the Wald ratio method was used to estimate the log odds change in AD risk for per standard deviation (SD) increment of circulating protein levels as proxied by the instrumental variables. The inverse-variance weighted (IVW) method was used to obtain the MR effects estimates for proteins with more than one instrument. The heterogeneity test was performed to assess the heterogeneity of the genetic instruments based on the Q statistic. We also performed additional analyses including simple mode, weighted mode, weighted median, and MR-Egger to account for horizontal pleiotropy. MR-Egger results were used only when the intercept indicated the presence of horizontal pleiotropy. High confidence AD protein changes in MR phase (HCPC-MR) for multiple tissues were defined as causally genetically predicted proteins associated with AD risk with FDR-adjusted P value < 0.05 as the significance level.

Bayesian colocalization test was leveraged to verify the causal associations between protein biomarkers and AD. To assess whether two associated signals (protein and AD risk) were consistent with a shared causal variant to distinguish the confounding of linkage disequilibrium, we employed summary statistics of proteins and AD meta-GWASs to perform Bayesian colocalization analysis based on the “coloc” package. The colocalization analysis included five hypotheses: (i) there was no causal variant for either protein or AD in the genomic locus (H0); (ii) there was one causal variant for protein only (H1); (iii) there was one causal variant for AD only (H2); there were two distinct causal variants for protein and AD (H3); (iv) there was a shared causal variant for protein and AD (H4). For each protein, we included SNPs within ±11Mb of the pQTL. When a protein had more than one pQTL, colocalization analysis was performed based on each pQTL, respectively, and the pQTL with the strongest evidence for colocalization was shown. The posterior probability for H4 (PP4) that was higher than 80% was considered strong evidence of colocalization. Since GWAS full summary statistics data for brain and CSF pQTLs was not publicly accessed in previous studies, the colocalization analysis was only performed for the plasma proteins.

### Comparison of protein changes across multiple human tissues

The comparisons of AD protein changes across brain bulk, CSF, and blood tissues were restricted to include the tissue-specific high-confidence AD candidates only. We performed a parallel comparison based on results from both SR and MR phases. Protein changes were categorized into (1) consistently increased/decreased in two or more tissues, (2) uniquely altered in brain bulk or CSF or blood tissues, (3) altered in the opposite direction in comparison of two or three tissues.

### Pathway and network analysis based on AD high confidence proteins

For HCPC-SR and HCPC-MR, we conducted Gene Ontology (GO) enrichment analyses using clusterProfiler, to determine which biological functions or processes were significantly enriched based on hypergeometric tests. Enriched GO pathways were determined based on a cut-off criterion of q-value ≤ 0.05. Protein–protein interaction (PPI) networks were generated in the STRING database (https://string-db.org/). The networks were edited in Cytoscape v3.10.0 and Adobe Illustrator. Pathway collections were annotated manually based on string GO enrichment outputs. We then used HCPC-SR to construct the PPI network, with the node size reflecting the AD candidate score (repeated counts of DEPs in previous proteomic studies).

### Exploratory study in Tear proteomics

#### Participant selection

For tear proteomics exploratory phase, a matched case-control study of 79 Chinese community-based older adults aged ≥60 years were enrolled, including 11 dementia, 34 mild cognitive impairment (MCI), and 34 age-, sex- and education-matched normal cognition controls (NC) (**Figure 1C**). The stages of dementia were determined based on the Clinical Dementia Rating (CDR) instrument, with a score ranging from 0 to 3 [37]. The Montreal Cognitive Assessment (MoCA) 5-minute tool was used for MCI screening [38], with a score ranging from 0 to 15. We defined dementia and MCI cases as the CDR score of ≥1 and 0.5, respectively. Individuals with the CDR score of 0 was defined as NC.

#### Tear sample collection and storage

Tear samples were obtained absorption of tear fluid in a paper Schirmer tear test strip (5mm x 30mm, Tianjin Jingming New Technology Development Co., Ltd., China). Tear samples from right and left eyes were collected in separate Schirmer strips and then store in aliquots at -80 °C. To avoid the within-subject biological differences between both eyes, we randomly selected the tear sample from the right eye for further proteomic analysis.

#### Peptide preparation

Tissue samples were processed using pressure cycling technology (PCT) for lysis and digestion. Initially, tissues were lysed with a buffer containing 6 M urea (Sigma) and 2 M thiourea (Sigma) in 100 mM ammonium bicarbonate. The resulting lysates were then digested with LysC and trypsin. Following digestion, peptides were acidified to pH 2–3 with trifluoroacetic acid and purified using C18 spin columns (17–170 μg capacity, Part No. HEM S18V, MA) prior to mass spectrometry analysis.

#### Data Acquisition with Mass Spectrometry

The PulseDIA acquisition was performed on a nanoflow DIONEX UltiMate 3000 RSLCnano System (Thermo Fisher Scientific, San Jose) coupled to a Q Exactive HF-X hybrid Quadrupole-Orbitrap (Thermo Fisher Scientific, San Jose). For each PulseDIA acquisition, 0.5 μg of peptides was injected and separated across a 30 min LC gradient (from 3 to 28% buffer B) at a flowrate of 6 μL/min (precolumn, 3 μm, 100 Å, 20 mm × 75 μm i.d.; analytical column, 1.9 μm, 120 Å, 150 mm × 75 μm i.d.). The total time for reequilibration and sample loading was about 30 min. False-discovery rate cut-offs for peptide and protein identifications were set at 1% for both. The detailed procedures are described in the previous studies[39].

#### Differential expression analysis in tear proteomic study

Based on the full proteomic data, we assessed whether the tissue-specific HCPC-SR and HCPC-MR can be quantified and identified in non-invasive tear proteomic study. We calculated the median abundance of certain candidates that replicated identified in tear, and compared the distribution across dementia, MCI, and NC stages. Multivariate analysis of covariance (MANCOVA), with age, sex, and educational level as covariates, was performed to identify differentially expressed proteins for comparisons between three dementia progression stages. Two-sided with P values <0.05 indicates statistical significance. Differentially expressed tear proteins for comparisons of interest (i.e., dementia vs. NC and MCI vs. and NC) were then presented as box plots. GO enrichment pathway analysis was conducted on significantly expressed tear proteins to explore the potential pathways. In the current study, all analyses were executed using R 4.4.1.

## Data Availability

Data are available upon reasonable request by directly contacting the corresponding author.

## Acknowledgements

We acknowledge the participants of tear exploratory study. We also acknowledge the staff for patient recruitment and data collection supports.

## Author’s contributions

Conceptualization, C.Y., J.S. (Jie Shen), and Y.H.; methodology, J.S. (Jie Shen) and Y.H.; systematic review, J.S. (Jie Shen) and M.W.; data extraction, J.S. (Jie Shen), M.W., and M.Y.; formal analysis, (Jie Shen) and Y.H.; investigation, (Jie Shen) and Y.H. and M.W.; Statistical support, X.L., J.S. (Jing Sun), and T.G.; writing—original draft preparation, J.S.; visualization, J.S.; writing—review and editing, C.Y., X.L. X.X., P.S., P.Y., X.Q., J.Y., H.C., T.G.; project administration, C.Y. All authors have read and agreed to the published version of the manuscript.

## Declarations

### Ethics approval and consent to participate

The study was approved by the research ethics committees of the Institutional Review Board of School of Public Health, Zhejiang University (ZGL20230123-5) and was registered under ClinicalTrial.gov (ID: NCT05886114). All participants signed written informed consent forms. All methods were performed in accordance with the relevant guidelines and regulations.

### Consent for publication

Not applicable.

